# Differences between plasma and CSF p-tau181 and p-tau231 in early Alzheimer’s disease

**DOI:** 10.1101/2021.12.10.21267467

**Authors:** Andrea Pilotto, Marta Parigi, Giulio Bonzi, Beatrice Battaglio, Elisabetta Ferrari, Lorenza Mensi, Alberto Benussi, Salvatore Caratozzolo, Maura Cosseddu, Rosanna Turrone, Silvana Archetti, Nicholas J Ashton, Henrik Zetterberg, Silvia Giliani, Alessandro Padovani

**Affiliations:** Department of Clinical and Experimental Sciences, Neurology Unit, University of Brescia, Italy; A. Nocivelli Institute for Molecular Medicine Spedali Civili and Department of Molecular and Translational Medicine, University of Brescia, Brescia, Italy; III Laboratory of Analyses, Brescia Hospital, Brescia, Italy; Department of Psychiatry and Neurochemistry, Institute of Neuroscience & Physiology, The Sahlgrenska Academy at the University of Gothenburg, Mo□lndal, Sweden; Wallenberg Centre for Molecular and Translational Medicine, University of Gothenburg, Gothenburg, Sweden; King’s College London, Institute of Psychiatry, Psychology and Neuroscience, Maurice Wohl Institute Clinical Neuroscience Institute, London, UK; NIHR Biomedical Research Centre for Mental Health and Biomedical Research Unit for Dementia at South London and Maudsley NHS Foundation, London, UK; Clinical Neurochemistry Laboratory, Sahlgrenska University Hospital, Mo□lndal, Sweden; Department of Neurodegenerative Diseases, UCL Institute of Neurology, London, UK; UK Dementia Research Institute at UCL, London, UK; Hong Kong Center for Neurodegenerative Diseases, Clear Water Bay, Hong Kong, China

**Keywords:** Phosphorylated-tau, plasma biomarkers, Alzheimer, Cerebrospinal fluid

## Abstract

Plasma phosphorylated tau species have been recently proposed as peripheral markers of Alzheimer’s disease pathology. In this cross-sectional study incuding ninety-one subjects, plasma p-tau181 and p-tau231 levels were elevated in the early symptomatic stages of AD, with similar levels than those of CSF. Plasma p-tau231 and p-tau181 were strongly related to CSF tau and amyloid and exhibited a high accuracy – close to CSF p-tau231 and p-tau181 – to identify AD already in the early stage of the disease. The findings might support the use as diagnostic and prognostic peripheral AD biomarkers in both research and clinical settings.

## INTRODUCTION

The development of blood-based biomarkers for Alzheimer’s disease (AD) is pivotal for a rapid screening and diagnosis and for tracking disease progression reducing the costs and burden of CSF and imaging assessments. Currently, the most promising blood biomarkers for detecting AD are amyloid Aβ_42/40_ ratio, glial fibrillary acid protein (GFAP), neurofibrillary light chain (NfL) and the newly identified different phosphorylated tau species [1-3]. Despite the recent validation of these biomarkers in clinical cohorts of AD patients [4-6], it is not yet well known how plasma p-tau species levels change in the very early stages of AD and across different neurodegenerative conditions. Furthermore, it is still controversial how different p-tau species in CSF and blood reflect the same pathological process and how they correlate with tau and amyloid AD markers. Thus, our main aim was to compare the levels of p-tau181 and p-tau231 in plasma and CSF and evaluate whether the levels of plasma differentiated AD patients from controls and patients with other neurodegenerative disorders. We hypothesized a significant increase in plasma p-tau181 and p-tau231 already in the early stages of AD and a correlation with AD-related biomarkers such as CSF Tau and amyloid levels.

## METHODS

### Study Population

This cross-sectional study included consecutive patients with a clinical diagnosis of AD [7], dementia or prodromal Lewy bodies (DLB) [8] or early stages of frontotemporal dementia (FTD) [9] who underwent CSF assessment at the Neurology Unit of Brescia. All patients underwent routine blood analyses, magnetic resonance imaging (MRI), a standardized full cognitive and behavioral assessment, including the Mini-Mental State Examination (MMSE) and the Neuropsychiatric Inventory (NPI) and an evaluation of basic and instrumental activities of daily living (ADL) [10]. Patients with mild cognitive impairment (MCI) or mild dementia were classified based on NIA-AA criteria and a diagnosis of AD was carried out according to CSF biomarker profile (see next section). The following exclusion criteria were applied: (1) cognitive deficits or dementia not fulfilling clinical criteria for AD, DLB or FTD; (2) prominent cortical or subcortical infarcts or brain/iron accumulation at imaging; (3) other neurologic disorders or medical conditions potentially associated with cognitive deficits; (4) bipolar disorder, schizophrenia, history of drug or alcohol abuse or impulse control disorder; (5) recent traumatic events or acute fever/inflammation potentially influencing CSF and plasma biomarkers. For biomarkers comparison, a group of non-neurological controls (HC, n=26) were included. This study was approved by the local ethics committee (NP 1471, DMA, Brescia) and was in conformity with the Helsinki Declaration; informed consent was obtained from all participants.

#### Fluid Biomarkers

At enrollment, 3 milliliters of CSF from each participant were collected. Approximately 10□mL venous blood was additionally collected in plastic tubes containing sodium ethylenediaminetetraacetic acid (EDTA). Lumbar puncture was performed according to the standardized protocol of the outpatient clinic, from 09:00 to 11:00 in the morning, after clinical informed written consent was obtained. CSF was collected in sterile polypropylene tubes and gently mixed to avoid gradient effects. CSF was centrifugated and firstly processed for standard biochemical analyses, whereas two milliliters of CSF were stored in cryotubes at −80°C before biomarkers testing. Only patients with normal routine measures were included in further analyses. CSF concentrations were measured in duplicate by an ELISA test (Innotest Tau antigen kit and Innotest PHOSHO-TAU). Standard cut-off values for AD used by our laboratory are Aβ_42_ < 650 ng/L, p-tau > 60 pg/mL, t-tau > 400 pg/mL and p-tau/ Aβ_42_ ratio > 0.9. In the study, patients were classified according to clinical diagnosis, CSF Aβ_42_ < 650 pg/mL and p-tau/ Aβ_42_ ratio > 0.9 in AD and other neurodegenerative dementias (NDD) [11, 12]. The blood samples were centrifuged at 2000 x g at 4□°C for 8□min; plasma supernatant was collected, divided into aliquots, and frozen at −□80□°C until further use. CSF and plasma p-tau181 and p-tau231 were quantified by using Simoa® pTau-181 Advantage V2 Kit and Simoa® pTAU-231 Advantage Kit through semi-automated SR-X Ultra-Sensitive Biomarker Detection System. The lower limit of quantification (LLOQ) as reported by manufacturer is 0.085 pg/mL for p-tau181 and 1.23 pg/ml for p-tau231. For plasma use of Simoa® pTAU-231 Advantage Kit (assay used for CSF by manufacturer) we applied a 4-fold standard dilution and the same conditions applied for plasma p-tau181 application. Outliers were defined as subjects with values above more than 5 standard deviations of the mean of the group-separate analyses including and excluding outliers were performed.

### Patient classification and statistical analysis

Data are presented as mean ± standard deviation for continuous variables and number (%) for categorical variables. Clinical and demographic characteristics as well as cognitive assessments and CSF and plasma p-tau181 and p-tau231 comparisons between diagnostic groups were performed using the Kruskal-Wallis, Bonferroni -corrected post-hoc analyses. The discriminative power in predicting AD CSF pattern was separately evaluated for CSF and plasma p-tau181 and p-tau231 using an overall accuracy analysis using the area under the curve (AUC) of a receiver operating characteristic curve (ROC). Correlations between CSF and plasma biomarkers were evaluated by partial correlation analyses adjusted for the effect of age and sex.

## RESULTS

### Participants’ characteristics and diagnosis according to AD CSF pattern

Ninety-one subjects, including 65 patients with cognitive impairment and 26 age-matched controls entered the study. The clinical assessment and CSF AD markers allowed the classification of patients in AD (n=43, of which 22 MCI and 21 with mild dementia) and other neurodegenerative disorders (NDD n=21, including 15 DLB and 6 FTD subjects). Table 1 shows biochemical and clinical characteristics of patients and controls. The groups did not differ for age, sex distribution but for MMSE and CSF AD biomarker levels (Table 1).

**Table 1.** Clinical characteristics of included patients. **Abbreviations:** Aβ_42_, amyloid beta 1-42; ADD, Alzheimer’s disease with dementia; HC, healthy controls; MCI-AD, mild cognitive impairment due to Alzheimer’s disease; MMSE, Mini-Mental state Examination; mL, milliliters; NDD, non-Alzheimer neurodegenerative disorders; pg, picograms. ¶significant comparison HC vs MCI-AD ¥significant comparison HC vs ADD # significant comparison NDD vs MCI-AD ^ significant comparison NDD vs ADD °significant comparison HC vs NDD

|  | HC (n=26) | NDD (n=22) | MCI-AD (n=22) | ADD (n=21) | p |
| --- | --- | --- | --- | --- | --- |
| Age, years | 65.0 ± 10.44 | 72.0 ± 4.3 | 69.6 ± 8.7 | 69.1 ± 8.3 | 0.28 |
| Disease duration, y | - | 2.6 ± 2.3 | 2.3 ± 2.5 | 2.4 ± 1.6 | 0.21 |
| Gender, F % (n) | 53% (14) | 36% (8) | 60% (26) | 60% (26) | 0.005 |
| MMSE | 29.3 ± 1.2 | 25.5 ± 4.2 | 26.1 ± 1.7 | 20.7 ± 1.8 | <0.001°¶# |
| **CSF markers** |  |  |  |  |  |
| t-tau, pg/mL | 250.6 ± 89.6 | 364.7 ± 163.5 | 644.4 ± 344.1 | 699.3 ± 350.1 | <0.001¶#¥^ |
| p-tau, pg/mL | 32.1 ± 8.9 | 44.8 ± 17.5 | 90.1 ± 34.1 | 133.5 ± 81.7 | <0.001¶#¥^ |
| Aβ<sub>42</sub>, pg/mL | 1142 ± 286 | 1269.4 ± 545.3 | 527.0 ± 159.7 | 540.4 ± 126.6 | <0.001¶#¥^ |
| p-tau/ Aβ<sub>42</sub> ratio | 0.28 ± 0.03 | 0.42 ± 0.15 | 1.77 ± 0.56 | 2.7 ± 2.3 | <0.001¶#¥^ |
| **CSF p-tau markers** |  |  |  |  |  |
| p-tau181, pg/mL | 15.7 ± 13.5 | 28.8 ± 21.5 | 80.5 ± 57.0 | 108.5 ± 99.6 | <0.001¶#¥^ |
| p-tau231, pg/mL | 30.1 ± 36.1 | 66.1 ± 66.2 | 142.2 ± 67.7 | 262.0 ± 230.1 | <0.001¶#¥^ |
| **Plasma p-tau biomarkers** |  |  |  |  |  |
| p-tau181, pg/mL | 1.91 ± 1.06 | 1.9 ± 0.95 | 3.8 ± 1.33 | 3.6 ± 1.8 | <0.001¶#¥^ |
| p-tau231, pg/mL | 2.1 ± 1.19 | 3.0 ± 1.72 | 4.6 ± 2.04 | 5.4 ± 2.02 | <0.001¶#¥^ |

### Plasma/CSF biomarkers levels and discriminant analyses

Plasma and CSF p-tau181 and p-tau231 levels were significantly higher in either MCI-AD or ADD and differentiated both MCI-AD and ADD from HC and NDD (p<0.001 for all post-hoc analyses, Table1 and Figure 1). Five outliers were identified; they included one case (ADD) for CSF-ptau181, one subject (NDD) for plasma p-tau181, two subjects (NDD n=1 and ADD n=1) and one case (MCI-AD) for plasma -tau231. No differences in p-tau181 or p-tau231 CSF levels between MCI-AD and ADD were observed. No correlation was found between age and sex and both CSF and plasma p-tau181 and p-tau231 levels.

**Figure 1.**
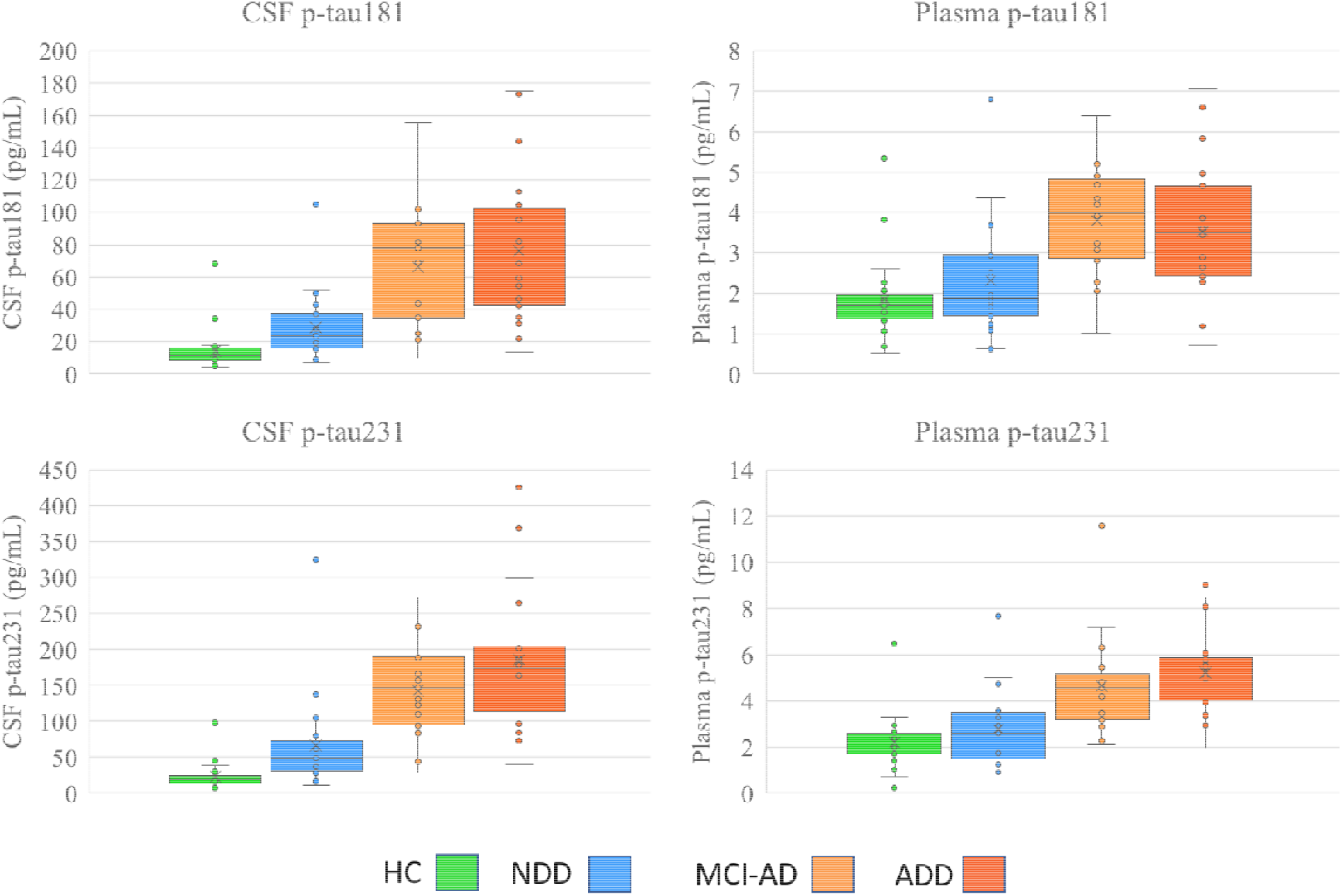
Differences in CSF and plasma p-tau181, p-tau231 levels according to each groups. Abbreviations: ADD, Alzheimer’s disease with dementia; HC, healthy controls; MCI-AD, mild cognitive impairment associated with AD pathology; NDD, non-Alzheimer neurodegenerative disorders.

We investigated how plasma and CSF p-tau181 and p-tau231 levels discriminate AD using ROC analysis. Plasma p-tau181 and p-tau231 as a biomarker accurately discriminated AD from non-AD individuals, with an AUC of 0.79 (CI95% 0.69-0.90%) and 0.87 (CI95% 0.79-0.95%), respectively. Similarly, CSF p-tau181 and p-tau231 had an AUC of 0.89 (0.82-0.96%) and 0.91 (IC 0.84-0.97%), respectively (Supplementary Figure 1).

### Correlations between plasma and CSF biomarkers

Plasma p-tau181 levels exhibited a strong correlation with CSF levels (r=0.44, p<0.001). A similar correlation was observed between plasma and CSF p-tau231 (r=0.44, p<0.001). P-tau181 and p-tau231 levels strongly correlated either in CSF (r=0.84, p=0.001) or plasma (r=0.63, p<0.001).

Plasma p-tau181 and p-tau231 exhibited significant positive correlation with CSF t-tau (r=0.32, p=0.001, r=-0.40, p<0.001), p-tau (r=0.44, p=0.001 and r=0.46, p<0.001), p-tau/Aβ_42_ ratio (r=0.43, p<0.001and r=0.47, p<0.001) and exhibited a negative correlation with Aβ_42_ levels (r=-0.42, p=0.001 and r=-0.36, p=0.008) (Figure 2).

**Figure 2.**
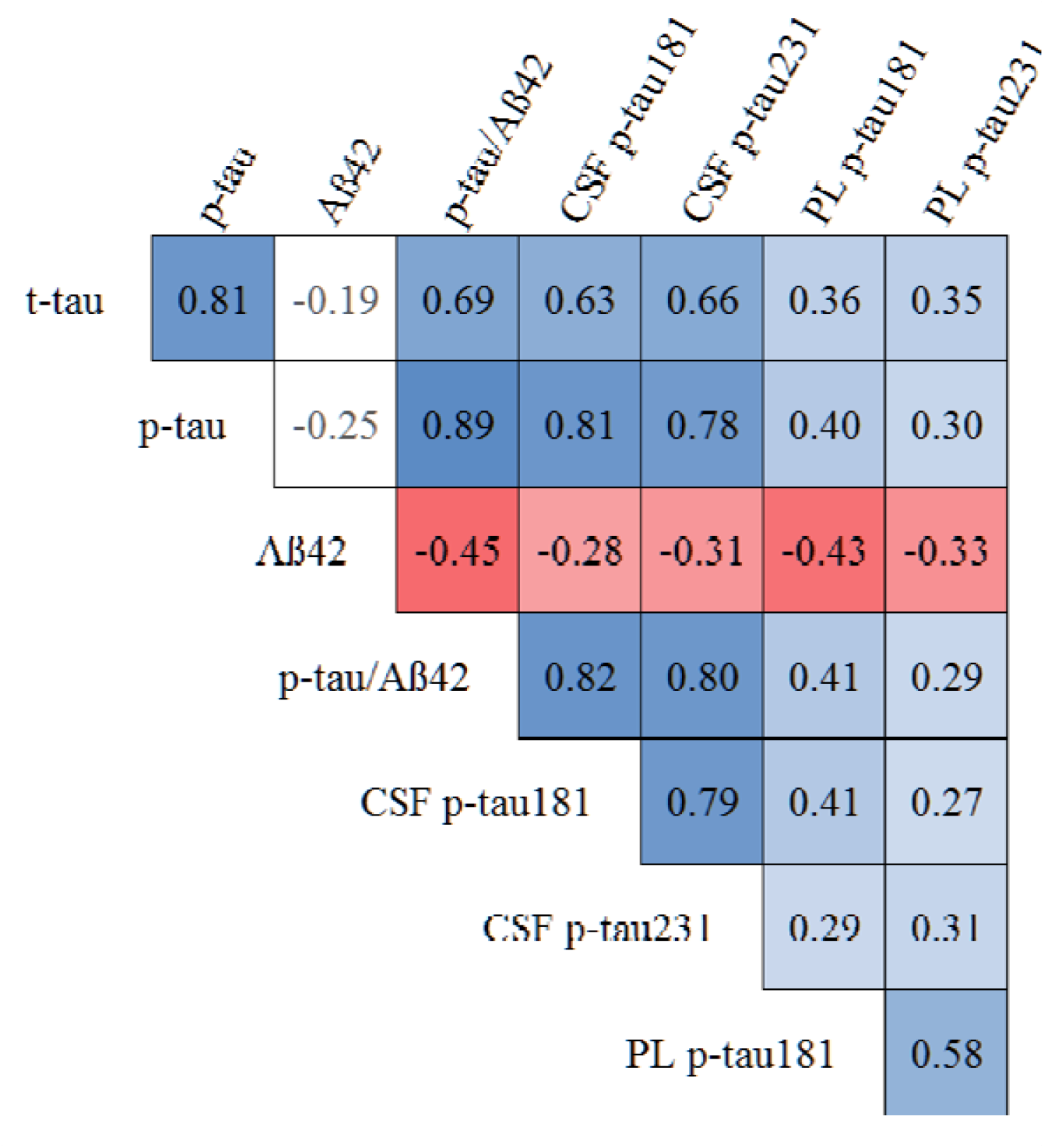
Correlations between CSF and plasma biomarkers in the sample. Blue indicates positive correlation, red indicates negative correlations, white indicates no significant correlations Abbreviations: Aβ_42_, amyloid beta 1-42; CSF, cerebrospinal fluid; PL, plasma

CSF p-tau181 and p-tau231 levels correlated positively with CSF t-tau (r=0.64, p=0.001; r=0.66, p=0.001), p-tau (r=0.84, p=0.001; r=0.81, p=0.001) and p-tau/ Aβ_42_ ratio (r=0.83, p=0.001; r=0.79, p=0.001) and negatively with Aβ_42_ (r=-0.29, p=0.03; r=-0.29, p=0.02 respectively) (Figure 2).

## DISCUSSION

In this study, plasma p-tau181 and plasma p-tau231 levels were significantly increased in early AD and mirrored CSF p-tau181 and p-tau231 levels. Further, there was a strong correlation between plasma and CSF levels of p-Tau181 and pTau231 with AD-related biomarkers including total tau, and Aβ_42_. Both plasma and CSF levels of p-tau181 and p-tau 231 shared high diagnostic accuracy for discriminate AD subjects from non-AD individuals.

These findings extend previous studies addressing blood-based biomarkers such as phosphorylated tau in independent cohorts [13-17]. Previous reports were particularly consistent for plasma p-tau181, as its levels are increased in AD [1, 3, 13, 18-22]. Very recently, Ashton and colleagues additionally identified plasma p-tau231 as a new potential marker of AD pathology claiming that it might be especially useful for the early phases of the disease [23].

In this study carried out on a series of subjects who underwent a standard clinical assessment and a CSF analysis of AD-related biomarkers for diagnostic purposes including total-tau, p-tau and Aβ_42_ [7], we have found that both p-tau181 and p-tau231 were closely related since the early stages of AD either in plasma or in CSF. Findings showed a strong correlation between the two different phosphorylated species p-tau181 and p-tau231 both in CSF and in plasma and a significant increase of both biomarkers in MCI-AD and ADD patients. For both p-tau181 and p-tau231, we confirmed the strong correlation between CSF and plasma levels [1, 2, 22]. The diagnostic performances of plasma p-tau181 and p-tau231 were very high and very close to the performance of CSF assays. In fact, the present findings argue for the great potential of both p-tau181 and p-tau231 as highly sensitive markers of AD pathology already in the MCI stages of the disease. Furthermore, the strong correlations observed for p-tau181 and p-tau231 with CSF p-tau/ Aβ_42_ ratio as well as with Aβ_42_, tau and p-tau markers strongly support the claim that these biomarkers are highly promising proxies for tracking disease progression over time, for predicting amyloid burden in preclinical and prodromal AD as well as for evaluating the effect of anti-amyloid therapies. In fact, the recent advances in pharmacological and non-pharmacological strategies for the treatment of AD requires a rapid change in the way the specialists are able to detect and follow AD patients [4-6]. The major limitation of the study resides in the sample size of both AD patients and non-AD patients thus requiring further larger validations with longitudinal progression to better define the diagnostic accuracy and ideal cut-offs [1, 2, 13]. The data, however, are highly consistent with published larger series and demonstrated the high performance and consistency between CSF and plasma levels, even including cases with frozen biosamples evaluated during a time span of several months. The number of outliers was indeed relatively low in CSF and plasma, underlying the usefulness of methods even in large and less characterised samples such as screening tool. Further studies are still needed in order to evaluate the role of preanalytical or clinical confounders for disentangling cases with borderline or discordant p-tau values. A direct comparison with other plasma markers including p-tau217, Aβ_42_/_40_ and GFAP will be also pivotal in order to evaluate the best biomarker combination both for diagnostic and prognostic purposes.

In conclusion, this study showed that the assessment of plasma p-tau 181 and p-tau231 ls a valuable method for early diagnosis of AD closely mirroring the discriminative accuracy of CSF p-tau 181 and p-tau231 markers.

In this framework, plasma biomarkers represent a unique opportunity for clinicians, pharma industries and-potentially-to healthcare systems - to reduce the costs and burden of assessment. and improve the ability to diagnose and track the disease progression in the new AD era.

## Data Availability

the data are available upon reasonable request from the authors

## ACKNOWLEDGEMENTS

The authors thank the patients who participated to the study and the health personnel involved in the clinical assistance and care of patients. This study is supported by the Airalzh-AGYR2020 grant issued to A.B. The authors acknowledge the contribution of Fondazione A. Nocivelli.

