## supplementary figure 1 for "Differences between plasma and CSF p-tau181 and p-tau231 in early Alzheimer’s disease"

**Supplementary Figure 1** Discriminant power for CSF and plasma p-tau181, p-tau231 for AD diagnosis. Abbreviations: AD, Alzheimer’s disease, CSF, cerebrospinal fluid.

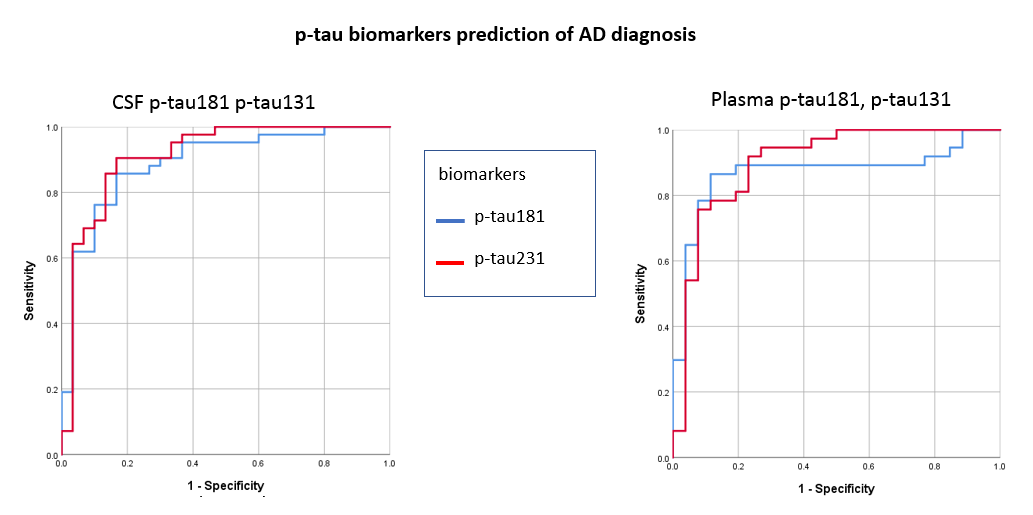
